# Chinese College Student Gamers Cohort (CCSGC): Multimodal Longitudinal Insights into Internet Gaming Disorder’s Biopsychosocial Mechanisms and Risk Trajectories

**DOI:** 10.64898/2026.04.01.26349949

**Authors:** Huang Yuchen, Zhou Guangdong, Liu Yifan, Xiang Shitong, Zheng Qihong, Wang Zifeng, Song Yixuan, Liu Wangyue, Wu Taoyu, Meng Shiqiu, Liao Yanhui, Jia Tianye, Shi Jie, Sun Yan

## Abstract

Internet gaming disorder (IGD) presents a significant public health challenge, yet its complex biopsychosocial mechanisms and dynamic risk trajectories remain poorly understood due to a scarcity of comprehensive longitudinal and multimodal cohorts. To address this critical gap, we established the Chinese College Student Gamers Cohort (CCSGC), a prospective, multimodal longitudinal study of 793 first-year undergraduates primarily playing Honor of Kings from 2022 Sept. The CCSGC integrates semi-annual psychosocial questionnaires, annual neuroimaging (EEG/fMRI), and biospecimen collection over multiple years. Baseline data revealed individuals with IGD (n=211) exhibited significantly higher gaming craving, psychological distress (depression, anxiety), impulsivity, and maladaptive motivational features compared to non-IGD gamers (regular players (RP) n=400; casual players (CP) n=182). Longitudinal analyses across four waves indicated bidirectional temporal associations between IGD severity and mental symptoms, and a stabilization of IGD incidence after an initial decrease. Furthermore, specific neurophysiological (e.g., N400 amplitude to game cues) and neuroimaging (e.g., superior parietal activation) markers were identified that correlated with IGD severity and predicted one-year outcomes in gaming disorder or social functioning. The CCSGC provides an invaluable resource for dissecting the heterogeneity, comorbidity, and intricate biopsychosocial mechanisms of IGD, holding significant potential to advance risk prediction, early identification, and targeted intervention strategies.

## Introduction

While digital gaming offers various cognitive and social benefits[1], a minority of players develop dysregulated engagement that can severely impact psychosocial functioning and mental well-being. Internet gaming disorder (IGD) is a globally recognized public health concern, characterized by persistent and recurrent gaming behavior leading to significant impairment or distress [2]. Recognizing its clinical significance, IGD has been classified as an addictive behavior in ICD-11 and proposed for further study in DSM-5[3, 4]. Despite rapidly expanding epidemiological evidence, a comprehensive understanding of IGD’s underlying biopsychosocial mechanisms and dynamic developmental trajectories remains critically limited[5, 6]. Specifically, robust longitudinal, multimodal investigations are scarce, hindering the identification of causal pathways, early markers, and modifiable targets essential for effective prevention and intervention strategies[7].

Existing research on IGD is predominantly cross-sectional, and while some longitudinal studies exist, they often suffer from short follow-up windows, infrequent assessment waves, and an over-reliance on self-reported symptoms, thereby restricting inferences about long-term course and the heterogeneous nature of the disorder [8, 9]. University students represent a particularly vulnerable population and a critical developmental window, transitioning into self-directed environments that may heighten the risk for problematic gaming[10]. In China, mobile games like Honor of Kings are immensely popular among this demographic, embedding team-based play and social network integration[11]. However, the specific biopsychosocial underpinnings and evolving risk profiles of IGD within this unique high-exposure group remain largely underexplored through integrated, multi-dimensional approaches.

Established risk factors for gaming disorder encompass a range of demographic, psychological, domains, including male sex, high impulsivity, low self-regulation, adverse childhood experiences, and familial or peer-related influences[12–15]. The IGD is frequently accompanied by a spectrum of behavioral problems, such as low academic performance, social isolation, and neglect of occupational or daily life responsibilities[16, 17]. Moreover, gaming disorder exhibits complex and often bidirectional associations with a broad array of psychiatric symptoms which most notably depression, anxiety disorders, and attention-deficit/hyperactivity disorder — suggesting overlapping neurobiological substrates and shared etiological pathways[18–20]. Despite accumulating cross-sectional evidence, the field currently lacks systematic longitudinal investigations, which are essential for disentangling causal relationships between these comorbidities and risk mechanisms.

A comprehensive understanding of IGD necessitates the exploration of its underlying neurobiological mechanisms, and EEG/ERP and fMRI studies have reported aberrant neural responses—including heightened cue-reactivity in reward-related circuits and deficits in cognitive control networks[6, 21]. A recent coordinate-based meta-analysis integrating structural and functional magnetic resonance imaging (MRI) evidence suggests convergent alterations in cingulate–prefrontal control regions and the supplementary motor area, alongside task-dependent functional abnormalities involving regions such as the insula and precuneus[22]. In parallel, a large rs-fMRI clustering analysis revealed substantial heterogeneity within IGD, characterized by largely different connectivity patterns across addiction-relevant networks [23]. Resting-state electroencephalography (EEG) studies also showed atypical oscillatory activity patterns in gaming disorder and closely related internet-addiction phenotypes, supporting the neurophysiological markers may complement MRI in characterizing addiction-relevant brain dynamics[24]. However, it is still unclear whether IGD encompasses heterogeneous neurobiological subtypes characterized by distinct harm mechanisms, and whether symptom severity and its associated neural signatures vary dynamically over time in heterogeneous patterns[8]. Longitudinal, multimodal approaches that repeatedly track neurophysiological and neuroimaging markers alongside clinical symptoms and key psychological features (such as motivation) are therefore needed to disentangle these complex interactions across IGD development[25].

To decisively address these significant limitations and advance the field’s understanding, we established the Chinese College Student Gamers Cohort (CCSGC) in September 2022. The CCSGC is a large-scale, prospective, multimodal longitudinal cohort comprising first-year undergraduates who are active players of Honor of Kings. This cohort is meticulously designed to provide unprecedented depth and breadth in tracking IGD development, integrating repeated psychosocial and behavioral phenotyping with nested EEG and imaging (fMRI), and biospecimen collection for genetic and epigenetic analyses. By capturing a broad continuum of gaming involvement, from casual players to individuals meeting DSM-5 IGD criteria, the CCSGC offers a powerful platform for mapping long-term individual trajectories, elucidating the complex biopsychosocial mechanisms of IGD, and predicting onset, persistence, and remission.

This article serves as the foundational introduction to the CCSGC. We detail the cohort’s comprehensive design, participant recruitment strategies, and extensive data collection protocols. Furthermore, we present key baseline characteristics of the cohort and highlight preliminary findings about IGD’s biopsychosocial abnormalities and longitudinal changes, that underscore the cohort’s methodological robustness and its potential for yielding novel insights. These initial data validate the CCSGC as a critical resource for mechanistic research, improving early identification, and informing evidence-based prevention and intervention strategies for IGD in college populations.

## Methods

### Study Design and Participant Recruitment

CCSGC is a prospective, multimodal, longitudinal study established in September 2022 at Tianjin Normal University. This cohort was designed to investigate the biopsychosocial mechanisms and developmental trajectories of Internet Gaming Disorder (IGD) among college students. Participants were recruited between October 2022 and October 2023 through mental health courses and university-affiliated social media platforms. Initial screening was conducted online for first-year undergraduates with experience playing the mobile game Honor of Kings. A total of 5,149 students completed the online screening questionnaire. After rigorous data cleaning, which excluded responses with implausibly short (< 180 s) or long (> 1 hour) completion times, incorrect attention-check answers, or indications of non-current gaming, a final baseline sample of 4,463 participants was retained. From this pool, 793 players, primarily engaged with Honor of Kings, were invited and consented to participate in the longitudinal follow-up study.

Participants were stratified into three groups based on baseline DSM-5 criteria for IGD and gaming behaviors: 1) IGD Group (n = 211): Met DSM-5 criteria (≥5 items) and averaged >2 hours of daily gaming. 2) Regular Players (RP, n = 400): Averaged >2 hours of daily gaming, considered gaming their primary online activity, but did not meet DSM-5 IGD criteria (<5 items).3) Casual Players (CP, n = 182): Did not consider gaming their primary online activity, averaged <1 hour of daily gaming, and did not meet DSM-5 IGD criteria.

All participants provided electronic informed consent, and the study was approved by the Biomedical Ethics Committees of Peking University (IRB00001052-23199) and the Ethics Committee of the Department of Psychology, Tianjin Normal University. The study was also registered with the China Clinical Trial Registration Center (HSR-26-000134). All raw data, preprocessing files, and processing documentation are being progressively uploaded to the Zenodo database (DOI: 10.5072/zenodo.475092) and will be made publicly available for research use.

### Baseline and Follow-up Procedures

At the baseline assessment, participants completed a comprehensive battery of electronic questionnaires administered via mobile devices in a classroom setting. The questionnaires covered demographic information, detailed gaming behaviors, gaming motivations, IGD symptoms based on DSM-5 criteria, and a range of mental health and psychological constructs. Following questionnaire completion, participants provided saliva samples using MF-728 M5 Hiper Saliva DNA Collection Tubes (2 ml) for subsequent genetic and epigenetic analysis. Participants who met the criteria for EEG and MRI scanning and provided voluntary consent were scheduled to undergo both EEG and MRI sessions. To avoid potential carryover effects of the tasks, participants who completed both modalities underwent the two scanning sessions at an interval of more than two weeks. Follow-up assessments are conducted semi-annually for questionnaire data and saliva collection, and annually for neuroimaging data. Consistent follow-up protocols, identical to baseline procedures, are maintained to ensure data consistency and comparability across waves. The cohort construction process is detailed in Figure 1

**Figure 1.**
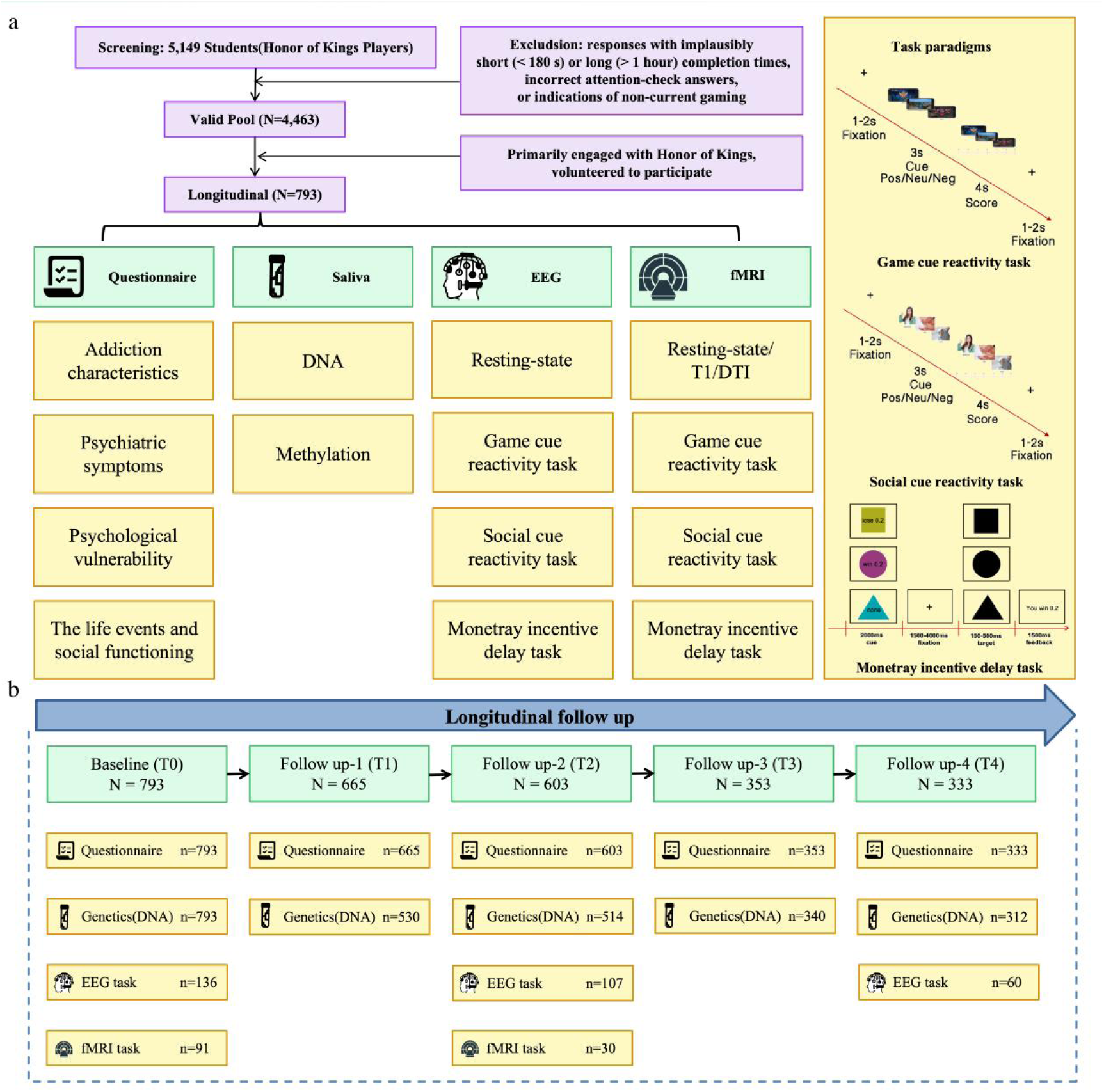
Flowchart. a. Cohort screening and enrollment criteria, and dimensions of data collection. b. Longitudinal follow-up procedures and sample sizes across time points and dimensions.

### Questionnaires

The questionnaires were organized into four thematic modules, please see Supplementary Table S1 for a complete list and references.

**1) Addiction characteristics:** Internet Addiction Test (IAT), DSM-5 (assessed by psychiatrically trained researchers following consensus training), Internet Gaming Disorder Scale-Short Form (IGDS9-SF), Visual Analogue Scale for Craving (VAS), Game Motivation Questionnaire, Fagerström Test for Nicotine Dependence (FTND) and Alcohol Use Disorders Identification Test (AUDIT).
**2) Psychological vulnerability:** the Chinese Big Five Personality Inventory-Brief Version, Brief Sensation Seeking Scale (BSSS), Barratt Impulsiveness Scale (Version 11) (BIS-11), Brief COPE Inventory, Parenting Styles and Dimensions Questionnaire (PSDQ), Childhood Trauma Questionnaire (CTQ), General Self-Efficacy Scale (GSES), Interpersonal Reactivity Index (IRI), and Sensitivity to Punishment and Sensitivity to Reward Questionnaire (SPSRQ).
**3) Psychiatric symptoms:** the Patient Health Questionnaire-9(PHQ-9), Generalized Anxiety Disorder 7-item Scale (GAD-7), Perceived Stress Scale (10-item) (PSS-10), Liebowitz Social Anxiety Scale (LSAS), Obsessive-Compulsive Inventory-Revised (OCI-R), Pittsburgh Sleep Quality Index (PSQI), and Reduced Morningness-Eveningness Questionnaire (MEQ).
**4) The life events and social functioning:** Evaluated via the Adolescent Self-Rating Life Events Checklist (ASLEC), Social Support Rating Scale (SSRS), Strengths and Difficulties Questionnaire (SDQ).

Except for the DSM-5 assessment, which was conducted by trained researchers, all other instruments were self-administered. Personality and psychological trait measures were collected only at baseline, whereas psychiatric symptoms and other longitudinal measures were collected at baseline and each follow-up visit (Table S1).

### Neuroimaging and EEG Sub-sampling and Eligibility

Participants from the longitudinal cohort were invited to participate in EEG or neuroimaging assessments based on voluntary interest and additional eligibility criteria. Inclusion criteria for EEG and neuroimaging required voluntary participation and current experience with Honor of Kings. Exclusion criteria included: 1) a diagnosis of any psychiatric or severe organic disease; 2) current use of psychotropic medications; 3) history of substance use disorders; 4) history of head injury or presence of intracranial metallic implants. Additional contraindications specific to EEG (e.g., allergy to conductive gel) and MRI (e.g., claustrophobia, standard MRI safety exclusions) were applied.

### Task Paradigms

Three computerized tasks were administered during EEG and fMRI sessions (Figure 1):

1. Game-Cue Reactivity Task: This task aimed to assess the neural and behavioral reactivity to gaming-related stimuli. Participants viewed and rated their craving (-3 to 3) for pictures depicting victory-related, defeat-related, and neutral game scenarios. Each trial consisted of a 3s picture/description presentation, a 4s rating period, and a 1-2s fixation interval, totaling 90 trials.
2. Social-Cue Reactivity Task: This task evaluated social reward processing and social sensitivity. Similar to the game-cue task, participants rated their pleasure (-3 to 3) in response to positive social, negative social, and personal neutral images.
3. Monetary Incentive Delay (MID) Task: Designed to assess reward and punishment sensitivity, this task presented cues indicating potential reward, punishment, or neutral outcomes, followed by a target requiring a rapid button press.

All visual stimuli used in the cue-reactivity tasks were independently pre-evaluated by a separate sample of gamers to ensure controlled arousal, valence, and familiarity (Supplementary Table S2).

### EEG Data Acquisition and Preprocessing

EEG data were acquired using a 32-channel Smarting Pro (Mbraintrain) system based on the 10-20 system. The recording included a 3-minute eyes-closed resting-state and task-state data during the three paradigms. Signals were amplified and sampled online at 500 Hz.

Preprocessing was performed using EEGLAB 9.0 in MATLAB 2022b. The pipeline included: 1) Re-referencing to linked mastoids. 2) Band-pass filtering (0.1-30 Hz) and a 50 Hz notch filter. 3) Independent Component Analysis (ICA) for artifact removal (ocular, cardiac). 4) Bad channel detection and spherical spline interpolation. 5) Epoching from -300 ms to +1000 ms relative to stimulus onset, with baseline correction. 6) Automated rejection of trials with amplitudes exceeding ±75 μV. 7). Event-Related Potential (ERP) components, specifically P300, N400, were identified based on grand-averaged waveforms and topographical maps. 8) Excluded for excessive artifacts or > 20% trial loss. Details on the eligibility rates were listed in the supplementary Table S3.

### MRI Data Acquisition and Preprocessing

Multimodal neuroimaging data were acquired using a Siemens 3T Prisma scanner equipped with a 64-channel head/neck coil at the Centre for MRI Research of Tianjin Normal University. The protocol included: 1) high-resolution T1-weighted structural images (MPRAGE sequence; TR/TI/TE = 2530/1100/2.98 ms; voxel size = 1.0 mm³) for anatomical reference; 2) resting-state fMRI (8-min acquisition; TR/TE = 2000/30 ms; 240 volumes; voxel size = 3.5 mm³) during which participants maintained eyes-closed wakefulness; 3) task-based fMRI using identical parameters during game-cue reactivity, social-cue reactivity, and monetary incentive delay paradigms; and 4) diffusion tensor imaging (DTI; TR/TE = 4400/68 ms; b-values = 1000/3000 s/mm²; 64 directions) for white matter microstructure assessment. Field maps were acquired for geometric distortion correction.

Data preprocessing followed standardized pipelines using SPM12. Functional MRI preprocessing included slice-time correction, rigid-body motion realignment, co-registration to T1-weighted images, spatial normalization to MNI space (resampled to 2 mm³ voxels), and smoothing with a 6-mm FWHM Gaussian kernel. Participants exhibiting excessive head motion (> 2.0 mm translation or > 2.0° rotation) were excluded from analysis. DTI preprocessing incorporated eddy-current correction and tensor fitting. These rigorous preprocessing steps ensured robust data quality for subsequent neural correlates analysis. Details on the eligibility rates were listed in the supplementary Table S3.

### DNA Extraction and Processing of Saliva Sample

Genetic data analysis aimed to explore associations between genetics and behavioral/brain functional measures, and the feasibility of genetic indicators as biomarkers. DNA extraction from saliva samples ensured sufficient quality (DNA concentration > 50 ng/µL, total quantity > 1 µg, and clear electrophoretic bands). Genotypes were obtained using the Infinium Asian Screening Array (ASA). Quality control was performed using PLINK v1.90, removing SNPs with call rate < 95%, minor allele frequency < 0.1%, and Hardy-Weinberg equilibrium P < 10⁻⁶. Samples with call rate < 95%, deviating ±3 standard deviation from heterozygosity rate mean, or proportion identity by descent PI_HAT > 0.2 were excluded. A total of 655 participants and 520,777 SNPs were retained after quality control. While methylation assays are planned for future work, genetic data are not currently scheduled for database submission or public release.

### Statistical Analysis

Model fit was assessed using the Tucker-Lewis Index (TLI), Root Mean Square Error of Approximation (RMSEA), and Bayesian Information Criterion (BIC). The associations between motivational dimensions and symptom scores were examined using Spearman’s correlation and Analysis of Variance (ANOVA). Generalized estimating equations (GEE) were used to evaluate the stability of changes in IGD incidence rates across multiple follow-up visits. Cross-lagged panel modeling was used to examine bidirectional causal relationships between different dimensions, particularly between gaming disorder and mental symptoms. False discovery rate (FDR) correction was applied for multiple comparisons.

## Results

### Cohort Characteristics and Follow-up Retention

The CCSGC enrolled 793 first-year undergraduates (mean age 19.07 ± 1.82 years; 32.4% male, 67.6% female) who primarily played Honor of Kings (Supplementary Table S4). This cohort includes individuals spanning a broad continuum of gaming involvement: 211 participants met DSM-5 diagnostic criteria for IGD, 400 were classified as regular players (RP), and 182 as casual players (CP). Robust retention rates were observed across the four follow-up waves for questionnaire and saliva collection (i.e., 83.9% for the first follow-up, 76.0% for the second, 44.5% for the third, 42.0% for the fourth), indicating sustained participant engagement (Supplementary Table S5).

### Baseline Differences among IGD, Regular and Casual Players

At baseline, the IGD group (n=211) demonstrated significantly higher gaming craving, addiction severity, and gaming hours compared to RP (n=400) and CP (n=182) (all p < 0.001). Factor analysis of gaming motivations identified four distinct subtypes: real-life social, online social, game reward, and escape. IGD participants scored highest in game reward and escape motivations (all p < 0.001), while RP/CP groups showed higher real-life social and online social motivations (all p < 0.05; Table S6, Figure S1). A higher proportion of males was observed in the IGD and RP groups (χ² = 35.52, p < 0.001)

In terms of psychological distress, IGD participants reported elevated depression (PHQ-9), anxiety (GAD-7), and perceived stress (PSS-10) (all p < 0.001). Furthermore, IGD was associated with a distinct psychobehavioral profile (Table 1), characterized by higher reward sensitivity, impulsivity, and neuroticism, increased childhood trauma experiences, and a greater likelihood of having other diagnosed mental disorders (all p < 0.001). Conversely, they exhibited lower self-efficacy, conscientiousness, extraversion, and poorer social functioning (negative social potency, diminished sociability) (all p < 0.05).

**Table 1.**
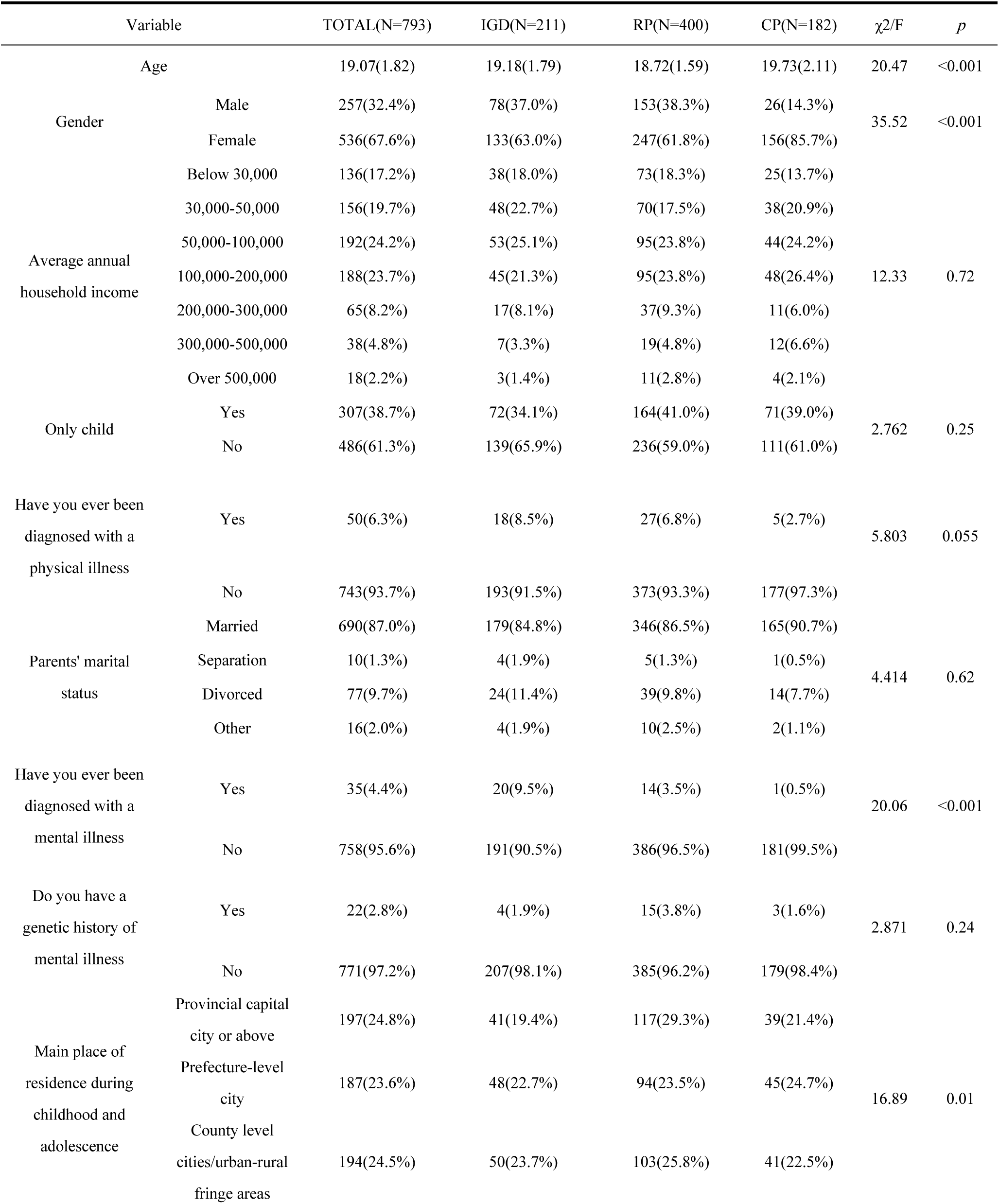
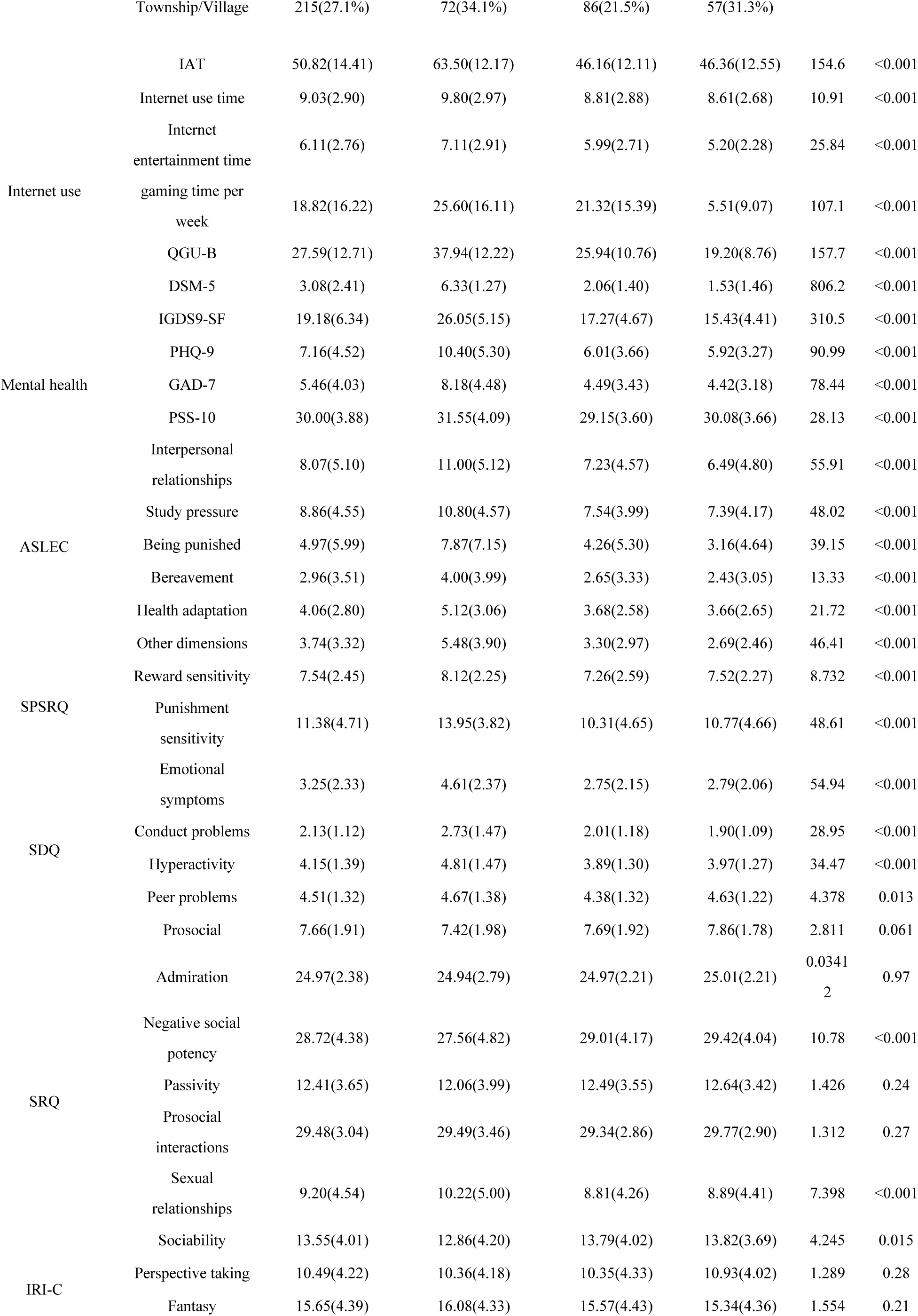
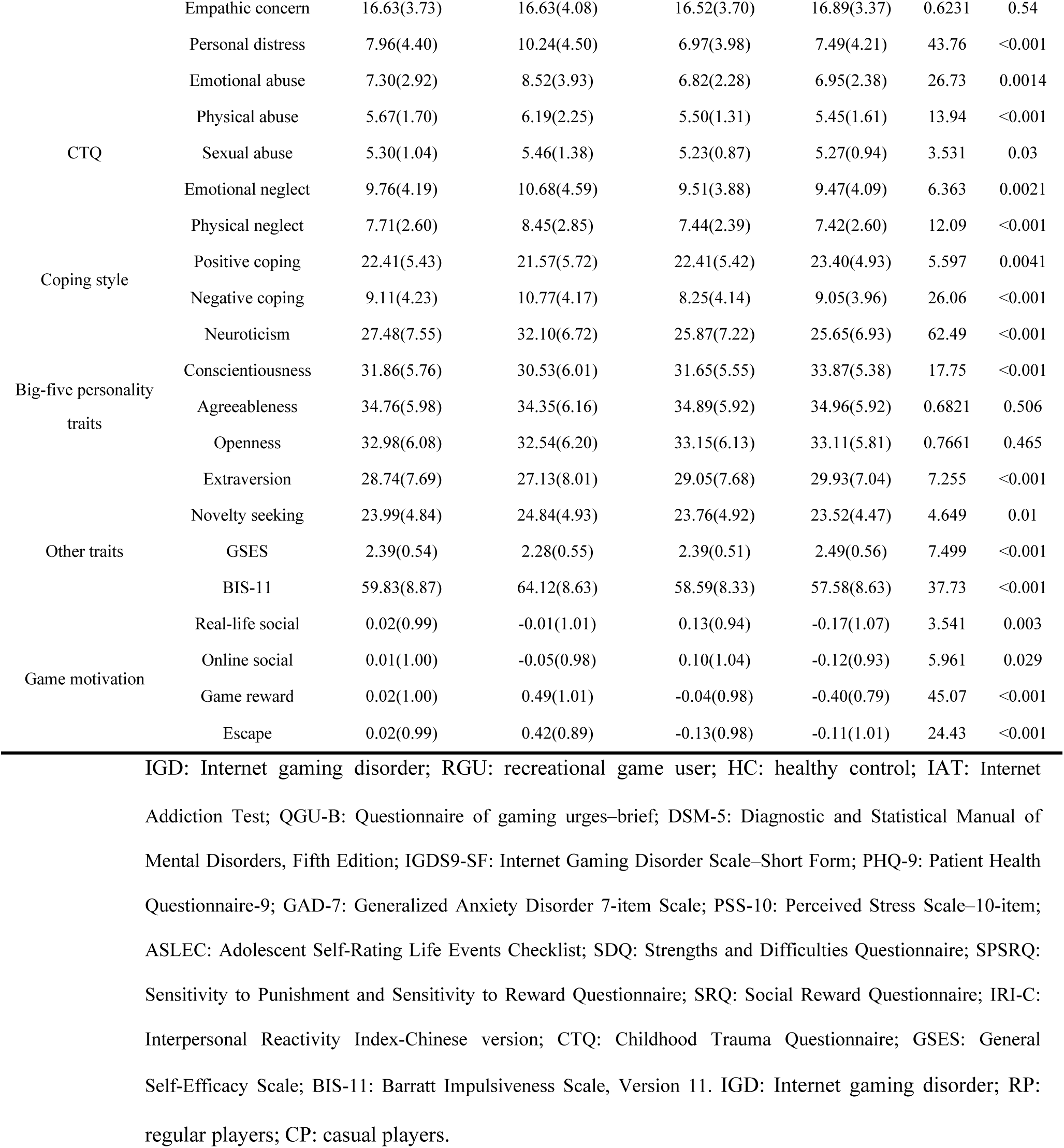
Demographic Characteristics of the Longitudinal Follow-up Cohort (N=793)

### Longitudinal Associations Between Gaming Disorder and Psychiatric Symptoms

Longitudinal analysis of IGD incidence across four follow-up assessments showed an initial significant decrease from baseline at the first follow-up (Wald χ² = 9.16, p = 0.0025), followed by stable rates that did not significantly differ from baseline in subsequent assessments (p > 0.05). Critically, cross-lagged panel modeling revealed a bidirectional causal association between gaming disorder severity module and psychiatric symptoms module (Baseline IGD to FU1 psychiatric symptoms: β = 0.20, p < 0.001; FU1 psychiatric symptoms to FU2 IGD: β = 0.18, p = 0.0086; Figure 2). No significant causal association was found between gaming disorder module and life events/social functioning module. Given the considerable temporal stability of psychological vulnerability, we did not examine its longitudinal associations with gaming disorder in the analyses.

**Figure 2.**
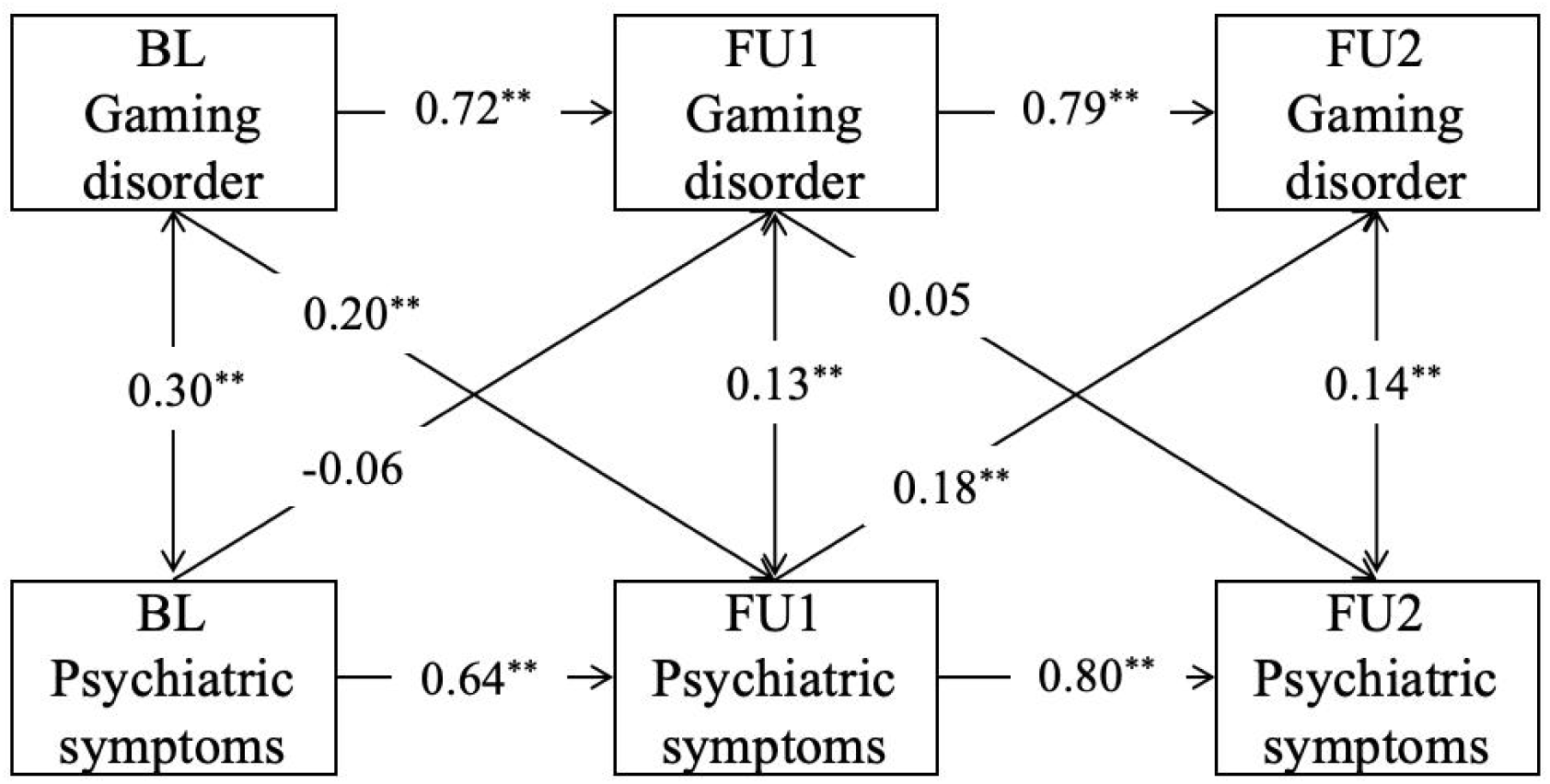
Cross-lag Analysis Between Gaming Disorder and Psychiatric Symptoms. BL: Baseline, FU1: Follow up 1, FU2: Follow up 2, **: *p* < 0.01.

### Neurophysiological Responses and Markers of IGD

Analysis of EEG data from 136 participants revealed distinct event-related potential (ERP) components in response to game and social cues (Figure 3; Supplementary Table S7). Specifically, P300 components were prominent over parieto-occipital regions, while N400 components were observed over frontal regions for both cue types.

**Figure 3.**
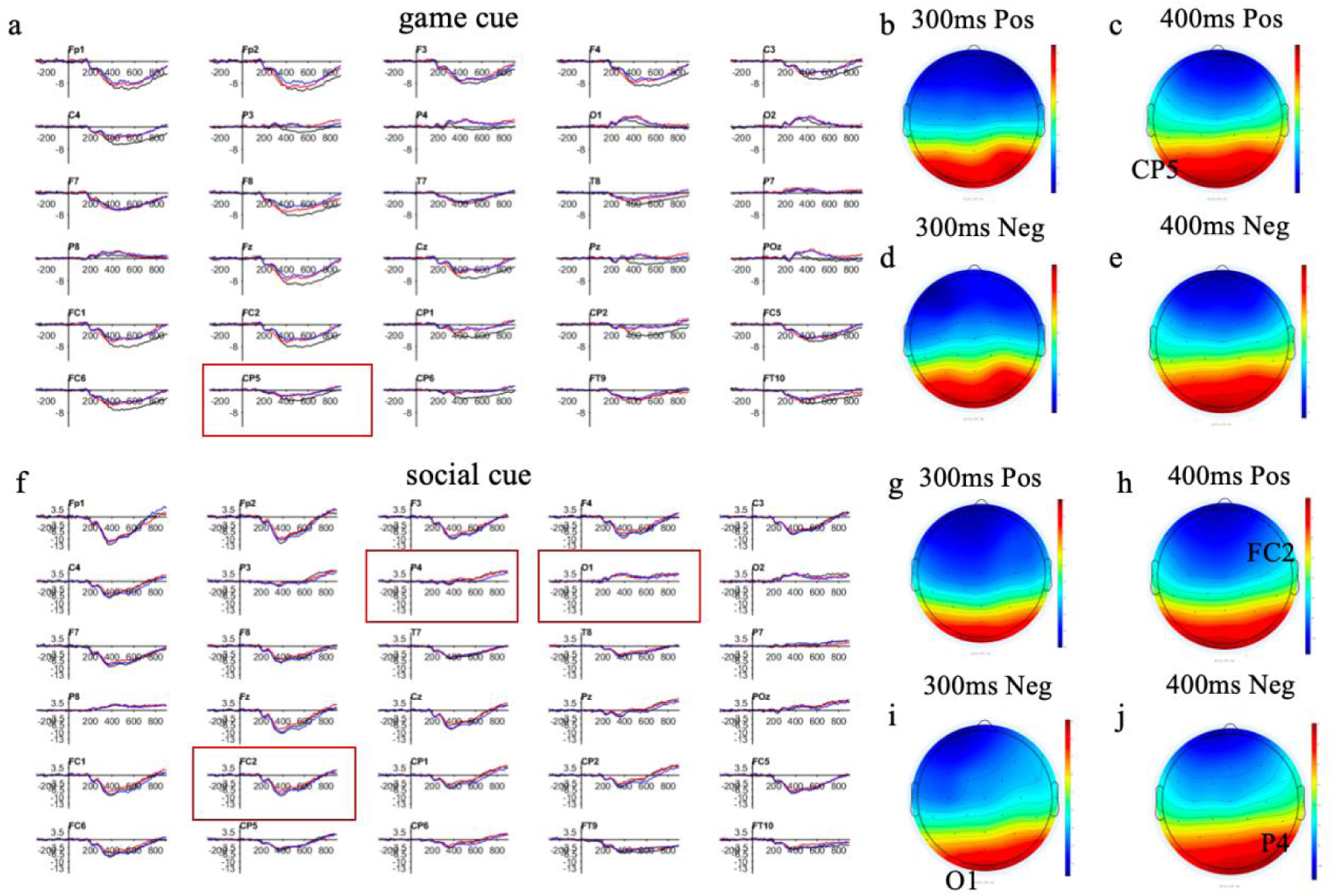
Grand-Average ERPs and Component Topographies during the Game Cue-Reactivity Task and Social Cue-Reactivity Task. a. Grand-average event-related potential (ERP) waveforms across 30 channels. Waveforms are depicted for neutral (black), positive (red), and negative (blue) conditions. b. Topographic map of EEG voltage distribution at 300 ms following the onset of positive game cues. c. Topographic map of EEG voltage distribution at 400 ms following the onset of positive game cues. d. Topographic map of EEG voltage distribution at 300 ms following the onset of negative game cues. e. Topographic map of EEG voltage distribution at 400 ms following the onset of negative game cues. f. Grand-average event-related potential (ERP) waveforms across 30 channels. Waveforms are depicted for neutral (black), positive (red), and negative (blue) conditions. g. Topographic map of EEG voltage distribution at 300 ms following the onset of positive social cues. h. Topographic map of EEG voltage distribution at 400 ms following the onset of positive social cues. i. Topographic map of EEG voltage distribution at 300 ms following the onset of negative social cues. j. Topographic map of EEG voltage distribution at 400 ms following the onset of negative social cues.

Regarding IGD-related electrophysiological markers, the mean N400 amplitude at CP5 in response to positive-neutral game cues was significantly negatively correlated with IGD severity (r = -0.23, p = 0.023; Figure 4a). Similarly, N400 amplitude at FC2 for positive-neutral social cues also negatively correlated with IGD severity (r = -0.28, p = 0.018; Figure 4b). These N400 components were uniquely associated with gaming disorder, not social-related behaviors. For negative social cues, O1 P300 amplitude correlated negatively with IGD severity (p = 0.041) and partially mediated the association between IGD severity and peer problems (indirect effect = 0.012, p = 0.020; Figure 4c). Furthermore, the N400 amplitude at CP5 (game cue) significantly predicted IGD severity one year later (r = -0.31, p = 0.028; Figure 4e), suggesting its stability as a neural marker.

**Figure 4.**
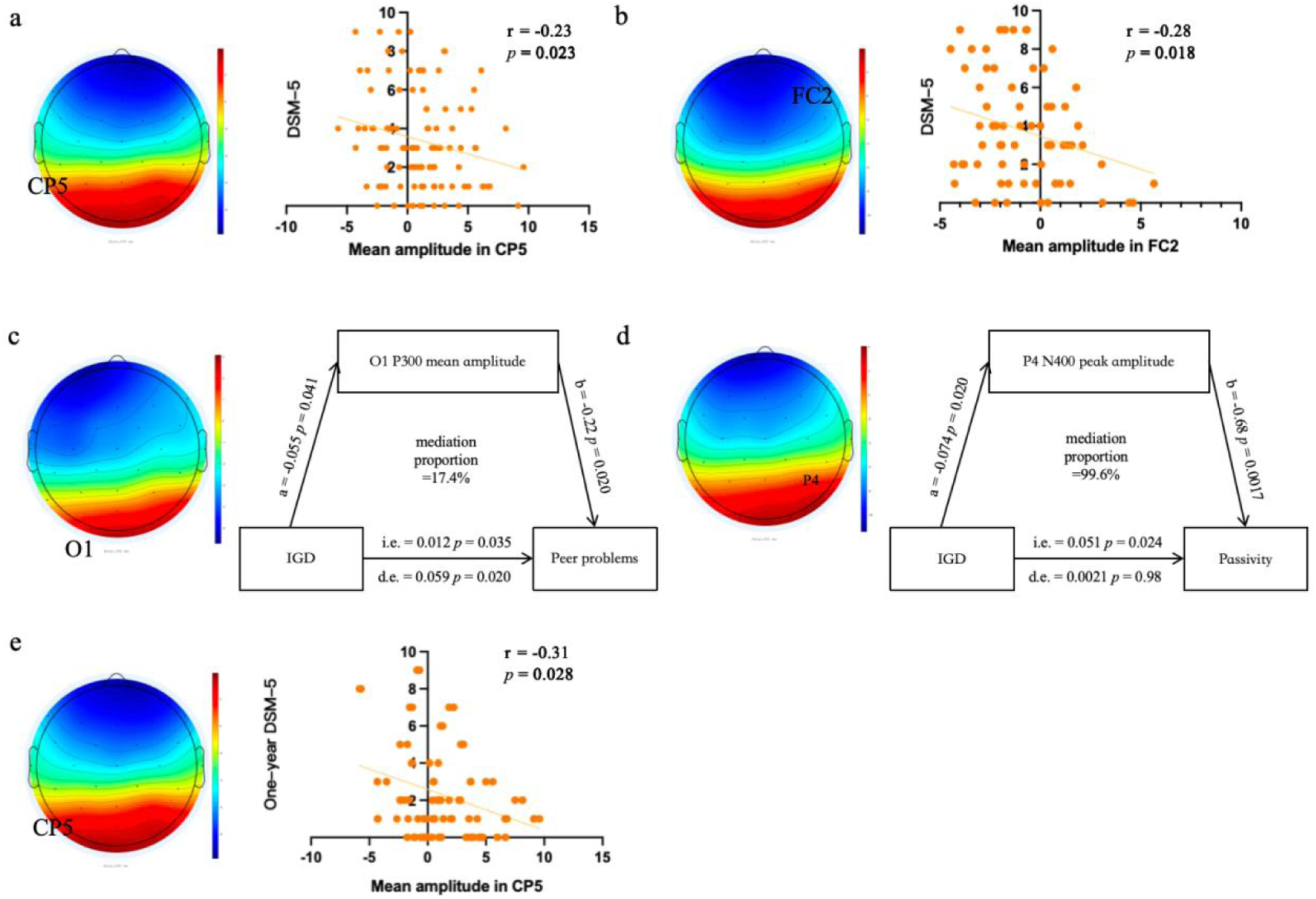
EEG Biomarkers of Gaming Disorder. a. Mean amplitude of the N400 component at the CP5 electrode for the positive-neutral condition in the game cue reactivity task. b. Mean amplitude of the N400 component at the FC2 electrode for the positive-neutral condition in the social cue reactivity task. c. Mean amplitude of the P300 component at the O1 electrode for the negative-neutral condition in the social cue reactivity task (i.e.=indirect effect, d.e.=direct effect). d. Peak amplitude of the N400 component at the P4 electrode for the negative-neutral condition in the social cue reactivity task (i.e.=indirect effect, d.e.=direct effect). e. The N400 mean amplitude at CP5 in the positive-neutral game cue condition predicted gaming disorder severity.

### Neuroimaging Activation Patterns and Markers of Gaming Disorder

Task-fMRI data from 91 participants revealed significant brain activation patterns during cue reactivity tasks (Figure 5; Supplementary Tables S7, S8). Game cues (positive-neutral contrast) engaged visual association cortices (e.g., lingual and fusiform gyri) and the precuneus, with concurrent decreases in specific frontal regions. Social cues similarly activated visual association cortex and the precuneus, alongside prefrontal regions.

**Figure 5.**
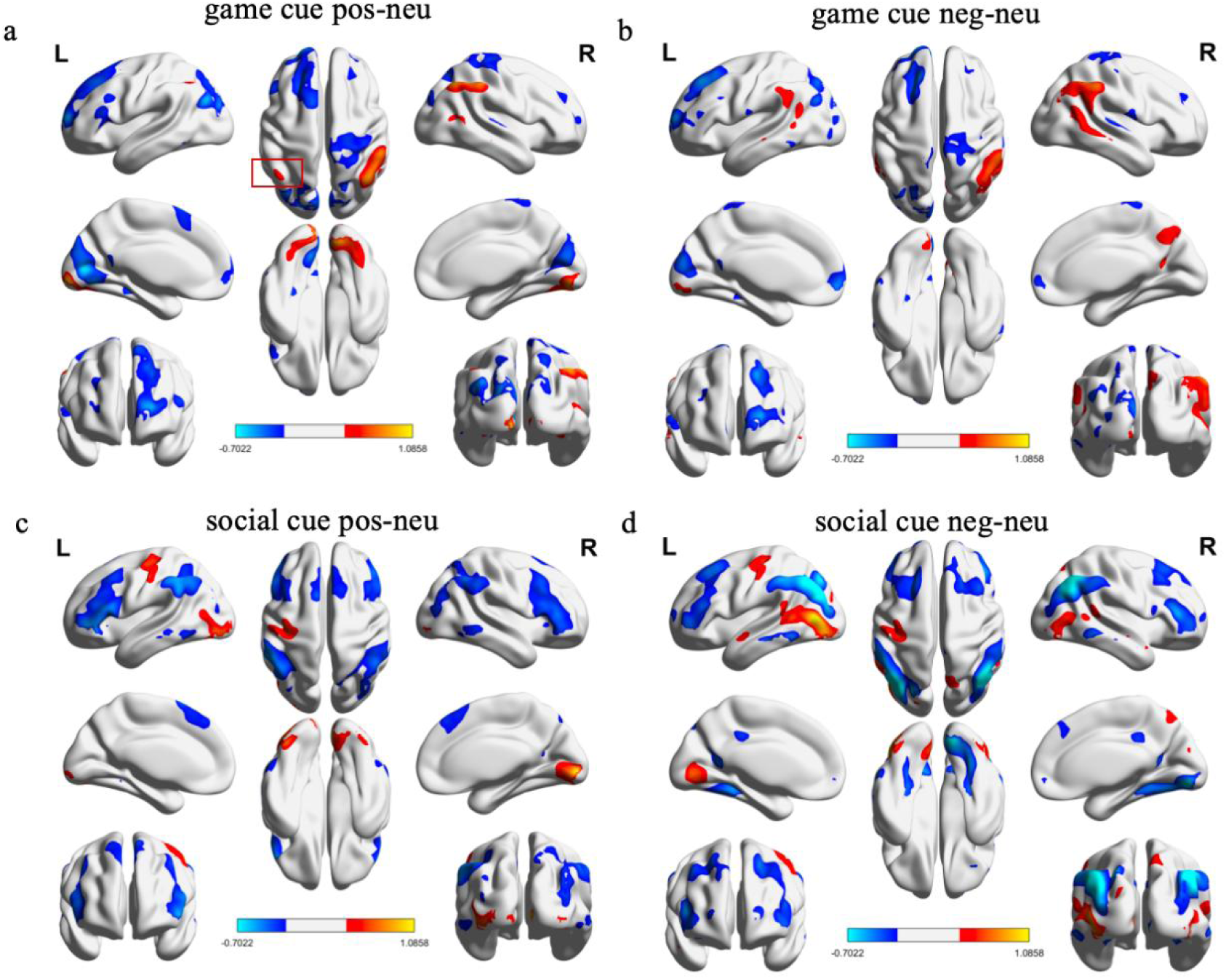
Brain Activation Pattern for Game Cues and Social Cues. a. Brain activation patterns for the game positive-neutral contrast. b. Brain activation patterns for the game negative-neutral contrast. c. Brain activation patterns for the social positive-neutral contrast. d. Brain activation patterns for the social negative-neutral contrast. Warm colors (red) indicate significant positive activation, while cool colors (blue) represent significant negative activation. All results are thresholded at a family-wise error (FWE) corrected significance level of *p* < 0.05.

After family-wise error (FWE) correction, activation in the left superior parietal gyrus during game positive-neutral cue reactivity was significantly and positively correlated with IGD severity (r = 0.32, p = 0.0023; Figure 6b). This superior parietal activation was also positively correlated with negative social potency (r = 0.37, p < 0.001; Figure 6c) and, importantly, predicted negative social potency one year later (r = 0.72, p < 0.001; Figure 6d). However, its correlation with IGD severity at one-year follow-up was not significantly positive (p = 0.11).

**Figure 6.**
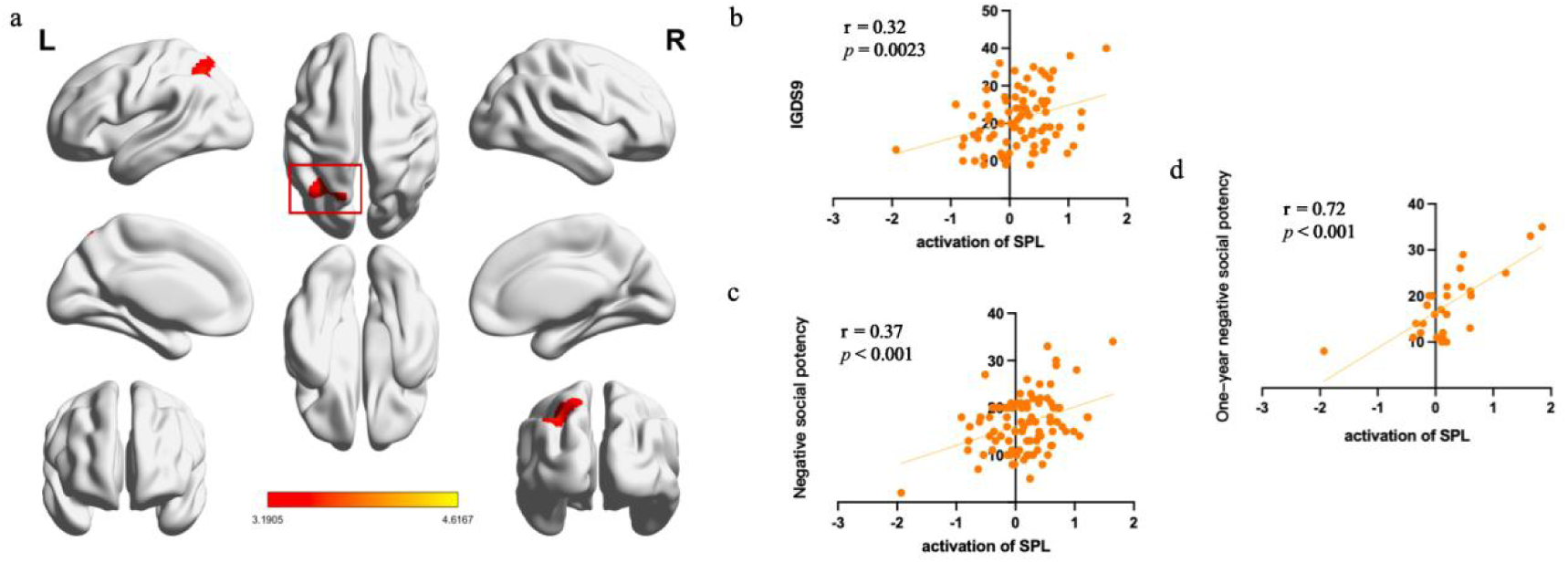
Neuroimaging Biomarkers of Gaming Disorder. a. Diagram depicting the left superior parietal lobule. b. Activation in the left superior parietal gyrus for the positive-neutral condition in the game cue reactivity task was associated with IGD. c. Activation in the left superior parietal gyrus for the positive-neutral condition in the game cue reactivity task was associated with negative social potency. d. Activation in the left superior parietal gyrus for the positive-neutral condition in the game cue reactivity task could predict one-year negative social potency.

## Discussion

The Chinese College Student Gamers Cohort (CCSGC) was meticulously designed as an enriched, deeply phenotyped, and multimodal longitudinal resource to investigate the complex biopsychosocial mechanisms and dynamic risk trajectories of Internet Gaming Disorder (IGD) in a crucial developmental window: first-year undergraduates who are active online gamers. To date, the cohort has successfully completed three years of follow-up, with ongoing longitudinal assessments encompassing semi-annual psychosocial evaluations, annual behavioral batteries, repeated electroencephalography (EEG) and functional MRI (fMRI) in a nested subsample, and biospecimen collection for genetic and epigenetic analyses. This paper presents the foundational design, comprehensive baseline characteristics, and compelling preliminary findings of the CCSGC, underscoring its methodological rigor and unique potential to advance our understanding of IGD.

Our baseline findings affirm the CCSGC’s value as a high-risk, high-information resource. We observed that IGD participants, representing a significant proportion (26.6%) of active gamers, exhibited markedly higher engagement, craving indices, and psychological distress (e.g., depression, anxiety) compared to recreational gamers. This aligns with and substantially extends meta-analytic evidence linking IGD to internalizing comorbidities and broader impairment severity [18]. Critically, IGD was associated with a distinct high-risk dispositional and developmental profile, characterized by elevated impulsivity, neuroticism, childhood trauma exposure, and maladaptive coping strategies, consistent with mechanistic models of IGD vulnerability [26, 27]. Beyond these established correlates, the CCSGC adds novel resolution by characterizing social-affective functioning, revealing elevated negative social potency and reduced sociability in IGD participants. These findings suggest that deficits in social connectedness may play a crucial role in differentiating heavy players who develop clinical impairment from those who remain recreational, thus pointing towards social resilience as a potential protective factor and a target for intervention [15, 28, 29]. Furthermore, our identification of four latent motivational dimensions (real-life social, online social, game reward, and escape) supports the view of IGD as a heterogeneous condition with diverse underlying pathways. This motivational heterogeneity underscores the need for personalized screening approaches that integrate motivational profiles beyond symptom duration alone.

The neurophysiological and neuroimaging results from the CCSGC provide critical insights into the neural mechanisms underlying altered cue processing in college gamers with IGD. At the EEG level, both game and social cues elicited robust P300 and N400 components, reflecting motivated attention and conflict monitoring [30]. Specifically, reduced N400 amplitude to positive game and social cues correlated with higher IGD severity, suggesting altered late-stage evaluative processing and meaning integration. This indicates that as IGD symptoms worsen, gaming and social cues may acquire increasingly consolidated motivational salience, potentially biasing the evaluation of real-world social signals [6, 31]. Notably, the left superior parietal activation to game cues was positively correlated with IGD severity and, crucially, predicted negative social potency one year later. This suggests that heightened attentional capture by rewarding game stimuli in more severe IGD may be linked to maladaptive social-affective outcomes, consistent with prior evidence for posterior parietal dysregulation during gaming-cue exposure [32]. This multimodal evidence for altered cue reactivity and its predictive power provides tangible neural markers for tracking IGD severity and progression.

Our longitudinal analyses reveal significant dynamics in IGD. We observed a bidirectional temporal association between IGD severity and mental symptoms, confirming their complex interplay and mutual influence over time, which has substantial implications for integrated treatment approaches. The observed stabilization of IGD incidence after an initial decrease from baseline provides valuable insights into the natural course of IGD in this population, suggesting a potential for spontaneous remission or adaptation in some individuals within the college environment. These longitudinal findings are crucial for developing dynamic models of IGD, moving beyond static cross-sectional correlations to uncover causal relationships and developmental trajectories.

The CCSGC represents a unique and powerful asset for behavioral addictions research, offering an unprecedented opportunity for multimodal, longitudinal investigation of IGD. Its strengths lie in its prospective design, large sample size of a vulnerable population (first-year college students), deep phenotyping across biological, psychological, and social domains, and the integration of advanced neuroimaging and genetic data. By focusing on a specific game (Honor of Kings), the cohort achieves high internal validity, enabling precise study of gaming-specific mechanisms. However, certain limitations warrant consideration. The primary recruitment from a single university in Tianjin introduces a potential regional bias, limiting generalizability to broader populations. Furthermore, while the cohort design enables multimodal integrative analyses, this inaugural paper primarily focuses on introducing the cohort and its initial findings; future work will fully leverage the integrated multi-omics and multi-modal neuroimaging data.

Looking ahead, the CCSGC will be instrumental in several promising research directions. We will continue to explore comorbidity patterns and their neurocognitive underpinnings to inform transdiagnostic perspectives on IGD. Crucially, the rich, multimodal, and longitudinal data will facilitate the development of advanced computational approaches for precise subtyping of IGD and improved diagnostic frameworks, moving beyond current categorical definitions. The data will also allow us to delineate the dynamic alterations in brain functional networks across different stages of the disorder and to utilize simultaneous EEG-fMRI for high spatiotemporal resolution investigation of cognitive and neural mechanisms underlying IGD onset and progression. Ultimately, the cohort will facilitate the study of complex interactions among biological predispositions, psychological characteristics, behavioral patterns, and environmental factors in shaping IGD trajectories.

## Conclusion

The CCSGC stands as a robust and continuously expanding multimodal longitudinal resource which characterized by its comprehensive design and initial three-year follow-up of 793 first-year undergraduates, has revealed significant insights into IGD. Our findings demonstrate that participants with IGD exhibit elevated mental symptoms, negative life events, childhood trauma, and distinct maladaptive motivational profiles at baseline. Longitudinally, cross-lagged analyses identified a bidirectional causal relationship between IGD and mental symptoms, while specific neurophysiological (e.g., N400/P300) and neuroimaging (e.g., superior parietal activation) markers showed predictive value for one-year outcomes in gaming disorder or social functioning. These results collectively underscore the profound heterogeneity, dynamic biopsychosocial mechanisms, and complex risk trajectories inherent in IGD. The CCSGC therefore holds immense promise for advancing mechanistic research, refining risk prediction models, informing early identification strategies, and developing targeted, evidence-based prevention and intervention programs. Crucially, it will also facilitate innovative computational approaches for precise subtyping and diagnosis, ultimately improving outcomes for college populations globally facing the challenges of IGD.

## Data Availability

All data produced in the present study are available upon reasonable request to the authors
All data produced in the present work are contained in the manuscript

## Acknowledgements

This work was supported by the STI2030-Major Projects (2022ZD0211200,2021ZD0201900), National Natural Science Foundation of China (NSFC, Grant No. 82471515 and 82271533).

