## supplementary materials for "Chinese College Student Gamers Cohort (CCSGC): Multimodal Longitudinal Insights into Internet Gaming Disorder’s Biopsychosocial Mechanisms and Risk Trajectories"

#### Table S1 Eligibility Rates for Assessment Questionnaires at Baseline and Follow-up Visits

| Questionnaire | Reference | Baseline | FU1 | FU2 | FU3 | FU4 |
|---|---|---|---|---|---|---|
| Internet Addiction Test (IAT) | Young, K. S. (1998). Internet addiction: The emergence of a new clinical disorder. *CyberPsychology & Behavior, 1*(3), 237–244. | × | × | × | × | × |
| Diagnostic and Statistical Manual of Mental Disorders, Fifth Edition – Internet Gaming Disorder (DSM-5 IGD) criteria | American Psychiatric Association. (2013). *Diagnostic and statistical manual of mental disorders* (5th ed.). American Psychiatric Publishing. | × | × | × | × | × |
| Internet Gaming Disorder Scale – Short Form (IGDS9-SF) | Pontes, H. M., & Griffiths, M. D. (2015). Measuring DSM-5 Internet gaming disorder: Development and validation of a short psychometric scale. *Computers in Human Behavior, 45,* 137-143. | × | × | × | × | × |
| Visual Analogue Scale for Craving (VAS) | Johanson, C. E., & Uhlenhuth, E. H. (1980). Drug preference and mood in humans: d-amphetamine. *Psychopharmacology, 71,* 275-279. | × | × | × | × | × |
| Liebowitz Social Anxiety Scale (LSAS) | Liebowitz, M. R. (1987). Social phobia. *Modern Problems of Pharmacopsychiatry, 22,* 141-173. | × | × | × | × | × |
| Fagerström Test for Nicotine Dependence (FTND) | Heatherton, T. F., Kozlowski, L. T., Frecker, R. C., & Fagerström, K. O. (1991). The Fagerström Test for Nicotine Dependence: A revision of the Fagerström Tolerance Questionnaire. *British Journal of Addiction, 86,* 1119-1127. | × | × | × | × | × |
| Alcohol Use Disorders Identification Test (AUDIT) | Saunders, J. B., Aasland, O. G., Babor, T. F., de la Fuente, J. R., & Grant, M. (1993). Development of the Alcohol Use Disorders Identification Test (AUDIT): WHO collaborative project on early detection of persons with harmful alcohol consumption—II. *Addiction, 88,* 791-804. | × | × | × | × | × |
| Patient Health Questionnaire-9 (PHQ-9) | Kroenke, K., Spitzer, R. L., & Williams, J. B. W. (2001). The PHQ-9: Validity of a brief depression severity measure. *Journal of General Internal Medicine, 16,* 606-613. | × | × | × | × | × |
| Generalized Anxiety Disorder 7-item Scale (GAD-7) | Spitzer, R. L., Kroenke, K., Williams, J. B. W., & Löwe, B. (2006). A brief measure for assessing generalized anxiety disorder: The GAD-7. *Archives of Internal Medicine, 166,* 1092-1097. | × | × | × | × | × |
| Chinese Big Five Personality Inventory – Brief Version (CBF-PI-B) | Wang, M. C., Dai, X. Y., & Yao, S. Q. (2011). Development of the Chinese Big Five Personality Inventory (CBF-PI) III: Psychometric properties of CBF-PI-B. *Chinese Journal of Clinical Psychology, 19,* 751-755. | × | — | — | — | — |
| Brief Sensation Seeking Scale (BSSS-8) | Hoyle, R. H., Stephenson, M. T., Palmgreen, P., Lorch, E. P., & Slater, M. D. (2002). Reliability and validity of a brief measure of sensation seeking. *Personality and Individual Differences, 32,* 401-414. | × | — | — | — | — |

| | | | | | | |
|---|---|---|---|---|---|---|
| Social Support Rating Scale (SSRS) | Xiao Shuiyuan (1994). The theoretical basis and research application of the Social Support Rating Scale Journal of Clinical Psychiatry, 4 (2), 3 | × | × | × | × | × |
| Barratt Impulsiveness Scale, Version 11 (BIS-11) | Patton, J. H., Stanford, M. S., & Barratt, E. S. (1995). Factor structure of the Barratt impulsiveness scale. *Journal of Clinical Psychology, 51,* 768-774. | × | — | — | — | — |
| Perceived Stress Scale – 10-item (PSS-10) | Cohen, S., Kamarck, T., & Mermelstein, R. (1983). A global measure of perceived stress. *Journal of Health and Social Behavior, 24,* 385-396. | × | × | × | × | × |
| Brief COPE Inventory | Carver, C. S. (1997). You want to measure coping but your protocol's too long: Consider the Brief COPE. *International Journal of Behavioral Medicine, 4,* 92-100. | × | — | — | — | — |
| The Parenting Styles and Dimensions Questionnaire (PSDQ) | Robinson, C. C., Mandleco, B., Olsen, S. F., & Hart, C. H. (2001). The Parenting Styles and Dimensions Questionnaire (PSDQ). In B. F. Perlmutter, J. Touliatos, & G. W. Holden (Eds.), Handbook of family measurement techniques: Vol. 3. Instruments & index (pp. 319 - 321). Thousand Oaks: Sage. | × | — | — | — | — |
| Childhood Trauma Questionnaire (CTQ) | Bernstein, D. P., & Fink, L. (1998). *Childhood Trauma Questionnaire: A retrospective self-report manual.* The Psychological Corporation. | × | — | — | — | — |
| Adolescent Self-Rating Life Events Checklist (ASLEC) | Liu Xianchen, Liu Lianqi, Yang Jie, etc Development and reliability and validity testing of adolescent life event scale [J]. Shandong Psychiatry, 1997, 10 (1): 15-19 | × | × | × | × | × |
| General Self-Efficacy Scale (GSES) | Schwarzer, R., & Jerusalem, M. (1995). Generalized Self-Efficacy scale. In J. Weinman, S. Wright, & M. Johnston (Eds.), *Measures in health psychology: A user's portfolio. Causal and control beliefs* (pp. 35-37). NFER-NELSON. | × | | | | |
| Sensitivity to Punishment and Sensitivity to Reward Questionnaire (SPSRQ) | Torrubia, R., Ávila, C., Moltó, J., & Caseras, X. (2001). The Sensitivity to Punishment and Sensitivity to Reward Questionnaire (SPSRQ) as a measure of Gray's anxiety and impulsivity dimensions. *Personality and Individual Differences, 31,* 837-862. | × | × | × | × | × |
| Strengths and Difficulties Questionnaire (SDQ) | Goodman, R. (1997). The Strengths and Difficulties Questionnaire: A research note. *Journal of Child Psychology and Psychiatry, 38,* 581-586. | × | × | × | × | × |
| Interpersonal Reactivity Index (IRI) | Davis, M. H. (1980). A multidimensional approach to individual differences in empathy. *JSAS Catalog of Selected Documents in Psychology, 10,* 85. | × | | | | |
| Obsessive-Compulsive Inventory – Revised (OCI-R) | Foa, E. B., Huppert, J. D., Leiberg, S., Langner, R., Kichic, R., Hajcak, G., & Salkovskis, P. M. (2002). The Obsessive-Compulsive Inventory: Development and validation of a revised version. *Psychological Assessment, 14,* 485-496. | × | × | × | × | × |
| Pittsburgh Sleep Quality Index (PSQI) | Buysse, D. J., Reynolds, C. F., Monk, T. H., Berman, S. R., & Kupfer, D. J. (1989). The Pittsburgh Sleep Quality Index: A new instrument | × | × | × | × | × |

| | | | | | | |
|---|---|---|---|---|---|---|
| | for psychiatric practice and research. *Psychiatry Research, 28,* 193-213. | | | | | |
| Reduced Morningness–Eveningness Questionnaire (MEQ-5) | Adan, A., & Almirall, H. (1991). Horne & Östberg Morningness-Eveningness Questionnaire: A reduced scale. *Personality and Individual Differences, 12,* 241-253. | × | × | × | × | × |

**Table S2 Valence Evaluations of Experimental Picture Cues Across Categories**

| Type | Picture | Arousal | Valence | Familiarity | Picture | Arousal | Valence | Familiarity |
|---|---|---|---|---|---|---|---|---|
| | game_1 | 2.40 | 1.15 | 4.00 | social_1 | 2.00 | 2.35 | 3.95 |
| | game_2 | 2.25 | 2.45 | 4.90 | social_2 | 2.35 | 1.10 | 3.15 |
| | game_3 | 2.20 | 1.05 | 3.10 | social_3 | 2.20 | 2.45 | 4.10 |
| | game_4 | 2.30 | 1.55 | 3.95 | social_4 | 2.60 | 2.20 | 4.85 |
| | game_5 | 2.25 | 1.75 | 3.35 | social_5 | 2.10 | 3.00 | 3.85 |
| | game_6 | 2.00 | 2.35 | 3.40 | social_6 | 2.25 | 2.60 | 4.05 |
| | game_7 | 2.00 | 1.85 | 4.50 | social_7 | 2.95 | 2.50 | 5.00 |
| | game_8 | 2.55 | 1.00 | 4.80 | social_8 | 2.25 | 1.40 | 4.95 |
| | game_9 | 2.50 | 2.30 | 3.85 | social_9 | 2.55 | 1.95 | 3.55 |
| | game_10 | 2.95 | 2.60 | 4.60 | social_10 | 3.00 | 2.45 | 3.30 |
| | game_11 | 2.35 | 1.75 | 4.90 | social_11 | 2.95 | 2.10 | 4.00 |
| | game_12 | 2.25 | 1.50 | 3.25 | social_12 | 2.20 | 2.35 | 3.25 |
| | game_13 | 2.75 | 1.70 | 3.00 | social_13 | 2.05 | 1.90 | 4.60 |
| | game_14 | 2.35 | 1.50 | 3.90 | social_14 | 3.00 | 1.60 | 4.85 |
| neutral | game_15 | 2.25 | 2.55 | 3.70 | social_15 | 2.35 | 2.10 | 3.80 |
| | game_16 | 2.45 | 1.80 | 4.55 | social_16 | 2.25 | 1.30 | 3.45 |
| | game_17 | 2.60 | 2.35 | 4.95 | social_17 | 2.00 | 2.45 | 4.85 |
| | game_18 | 2.15 | 1.30 | 4.60 | social_18 | 2.15 | 1.70 | 4.65 |
| | game_19 | 2.95 | 2.65 | 4.55 | social_19 | 3.00 | 1.40 | 3.15 |
| | game_20 | 2.40 | 2.20 | 3.90 | social_20 | 2.95 | 1.45 | 3.15 |
| | game_21 | 2.10 | 1.10 | 4.20 | social_21 | 2.55 | 1.70 | 4.05 |
| | game_22 | 2.85 | 1.50 | 4.35 | social_22 | 2.40 | 2.15 | 3.85 |
| | game_23 | 2.90 | 2.55 | 4.95 | social_23 | 2.80 | 1.05 | 3.35 |
| | game_24 | 3.00 | 2.05 | 3.05 | social_24 | 2.80 | 1.10 | 4.00 |
| | game_25 | 2.55 | 1.50 | 3.50 | social_25 | 2.10 | 2.25 | 3.40 |
| | game_26 | 2.20 | 1.40 | 4.50 | social_26 | 2.70 | 2.35 | 3.10 |
| | game_27 | 2.05 | 1.10 | 4.30 | social_27 | 2.80 | 2.00 | 4.25 |
| | game_28 | 2.00 | 2.65 | 3.40 | social_28 | 2.80 | 1.15 | 4.60 |
| | game_29 | 2.25 | 2.45 | 4.20 | social_29 | 2.70 | 1.80 | 3.85 |
| | game_30 | 2.45 | 2.85 | 3.35 | social_30 | 2.80 | 2.55 | 4.85 |
| | game_31 | 4.90 | 4.60 | 4.85 | social_31 | 4.90 | 3.45 | 4.00 |
| positive | game_32 | 3.40 | 4.45 | 3.45 | social_32 | 4.45 | 3.05 | 4.55 |
| | game_33 | 3.75 | 4.70 | 4.70 | social_33 | 4.30 | 3.60 | 4.90 |

| | | | | | | | | |
|---|---|---|---|---|---|---|---|---|
| | game_34 | 4.10 | 4.85 | 4.35 | social_34 | 3.10 | 3.00 | 4.40 |
| | game_35 | 4.55 | 4.95 | 4.80 | social_35 | 3.30 | 3.25 | 4.95 |
| | game_36 | 3.05 | 4.90 | 4.85 | social_36 | 4.25 | 4.70 | 4.05 |
| | game_37 | 3.15 | 4.10 | 3.95 | social_37 | 3.05 | 4.60 | 3.60 |
| | game_38 | 3.45 | 3.10 | 4.10 | social_38 | 4.65 | 3.40 | 3.15 |
| | game_39 | 3.45 | 3.65 | 3.10 | social_39 | 3.45 | 3.85 | 3.25 |
| | game_40 | 4.30 | 4.85 | 4.75 | social_40 | 3.70 | 3.30 | 3.55 |
| | game_41 | 4.55 | 4.30 | 3.30 | social_41 | 4.95 | 3.65 | 4.95 |
| | game_42 | 3.20 | 4.80 | 4.35 | social_42 | 4.20 | 4.55 | 4.50 |
| | game_43 | 3.80 | 4.95 | 3.15 | social_43 | 4.55 | 4.10 | 4.50 |
| | game_44 | 4.30 | 3.00 | 3.05 | social_44 | 3.50 | 4.85 | 4.70 |
| | game_45 | 3.65 | 3.70 | 4.85 | social_45 | 4.90 | 4.70 | 3.00 |
| | game_46 | 3.85 | 3.95 | 4.25 | social_46 | 4.95 | 4.45 | 4.40 |
| | game_47 | 4.70 | 4.05 | 4.60 | social_47 | 3.95 | 4.15 | 4.25 |
| | game_48 | 3.10 | 3.95 | 4.85 | social_48 | 4.05 | 3.15 | 3.50 |
| | game_49 | 4.10 | 3.40 | 3.95 | social_49 | 3.05 | 3.80 | 4.10 |
| | game_50 | 4.90 | 3.50 | 4.05 | social_50 | 4.25 | 4.10 | 3.30 |
| | game_51 | 4.15 | 4.20 | 4.60 | social_51 | 4.70 | 3.80 | 4.15 |
| | game_52 | 3.30 | 4.90 | 5.00 | social_52 | 3.20 | 3.95 | 4.85 |
| | game_53 | 3.95 | 3.45 | 5.00 | social_53 | 4.60 | 4.20 | 3.00 |
| | game_54 | 4.35 | 3.70 | 4.85 | social_54 | 3.95 | 3.95 | 3.00 |
| | game_55 | 4.90 | 4.45 | 4.65 | social_55 | 4.00 | 4.55 | 4.20 |
| | game_56 | 3.25 | 3.30 | 3.15 | social_56 | 3.85 | 3.35 | 4.30 |
| | game_57 | 4.30 | 4.20 | 3.40 | social_57 | 4.70 | 3.60 | 4.40 |
| | game_58 | 3.40 | 3.55 | 3.25 | social_58 | 3.85 | 4.10 | 3.80 |
| | game_59 | 4.75 | 3.85 | 4.10 | social_59 | 3.85 | 3.00 | 3.15 |
| | game_60 | 4.80 | 3.35 | 3.55 | social_60 | 4.35 | 3.75 | 4.65 |
| | game_61 | 3.20 | 4.65 | 4.40 | social_61 | 3.35 | 3.95 | 4.40 |
| | game_62 | 3.50 | 3.05 | 4.50 | social_62 | 4.80 | 4.25 | 3.70 |
| | game_63 | 3.50 | 3.60 | 3.30 | social_63 | 3.25 | 4.15 | 4.95 |
| | game_64 | 3.15 | 3.95 | 3.25 | social_64 | 3.05 | 4.00 | 3.55 |
| | game_65 | 4.10 | 4.90 | 3.15 | social_65 | 3.05 | 3.55 | 3.00 |
| | game_66 | 3.20 | 3.30 | 4.65 | social_66 | 4.20 | 4.55 | 4.30 |
| | game_67 | 3.85 | 3.30 | 4.50 | social_67 | 4.70 | 3.85 | 4.35 |
| negative | game_68 | 4.15 | 4.25 | 4.05 | social_68 | 3.10 | 3.15 | 4.95 |
| | game_69 | 3.75 | 4.90 | 4.60 | social_69 | 3.00 | 3.85 | 4.75 |
| | game_70 | 3.60 | 3.60 | 4.00 | social_70 | 3.95 | 4.75 | 4.80 |
| | game_71 | 3.30 | 3.75 | 5.00 | social_71 | 3.35 | 4.70 | 3.80 |
| | game_72 | 4.65 | 4.15 | 4.95 | social_72 | 3.40 | 4.45 | 3.60 |
| | game_73 | 5.00 | 3.00 | 4.00 | social_73 | 4.65 | 3.05 | 4.00 |
| | game_74 | 4.85 | 4.30 | 3.50 | social_74 | 4.70 | 4.85 | 4.05 |
| | game_75 | 3.75 | 3.65 | 3.85 | social_75 | 3.25 | 3.20 | 4.90 |
| | game_76 | 4.70 | 3.30 | 4.05 | social_76 | 4.00 | 4.85 | 4.90 |

| | | | | | | | |
|---|---|---|---|---|---|---|---|
| game_77 | 3.15 | 4.55 | 3.35 | social_77 | 3.00 | 4.55 | 4.80 |
| game_78 | 4.55 | 4.25 | 4.50 | social_78 | 3.25 | 3.90 | 3.20 |
| game_79 | 3.05 | 4.25 | 4.90 | social_79 | 3.25 | 3.10 | 4.25 |
| game_80 | 4.25 | 3.95 | 4.50 | social_80 | 3.85 | 4.75 | 4.40 |
| game_81 | 4.00 | 5.00 | 3.65 | social_81 | 4.50 | 3.15 | 3.10 |
| game_82 | 3.40 | 4.40 | 4.20 | social_82 | 3.55 | 3.65 | 3.20 |
| game_83 | 4.80 | 3.05 | 3.40 | social_83 | 3.00 | 3.05 | 5.00 |
| game_84 | 4.55 | 3.80 | 4.45 | social_84 | 4.10 | 4.05 | 4.90 |
| game_85 | 4.65 | 3.40 | 4.15 | social_85 | 4.55 | 4.35 | 3.70 |
| game_86 | 4.20 | 4.10 | 3.10 | social_86 | 3.75 | 4.75 | 4.65 |
| game_87 | 3.40 | 4.80 | 3.35 | social_87 | 3.40 | 4.95 | 3.95 |
| game_88 | 3.45 | 4.15 | 4.05 | social_88 | 3.20 | 3.50 | 3.40 |
| game_89 | 3.95 | 4.45 | 3.15 | social_89 | 3.40 | 4.65 | 3.55 |
| game_90 | 4.75 | 4.35 | 4.35 | social_90 | 3.50 | 3.90 | 3.45 |

**Table S3 Data Acquisition and Eligibility Rates Across Modalities**

| Multimodal data | Baseline (Eligible N/Total N) | FU1 (Eligible N/Total N) | FU2 (Eligible N/Total N) | FU3 (Eligible N/Total N) |
|---|---|---|---|---|
| EEG | 136/188 | 107/107 | 60/60 | — |
| fMRI | 91/99 | 30/30 | — | — |
| Saliva | 655/793 | 530/665 | 514/603 | 340/353 |

**Table S4 Demographic and Clinical Characteristics of the Overall Sample (N=4463)**

| Variable | | Mean(SD)/N(%) |
|---|---|---|
| Age | | 18.88(0.98) |
| Gender | Male | 2391(53.6%) |
| | Female | 2072(46.4%) |
| Internet use | Internet use time | 8.48(3.72) |
| | Internet entertainment time | 5.95(3.48) |
| | IAT | 49.55(14.92) |
| | IGDS9-SF | 20.74(7.29) |
| SRQ | Admiration | 24.38(3.42) |
| | Negative social potency | 28.19(5.57) |
| | Passivity | 12.65(4.05) |
| | Prosocial interactions | 29.13(4.25) |
| | Sexual relationships | 9.72(5.36) |
| | Sociability | 13.72(4.46) |

IAT: Internet Addiction Test; IGDS9-SF: Internet Gaming Disorder Scale–Short Form; SRQ: Social Reward Questionnaire.

### Table S5 Retention Rates across Follow-up Time Points in the Longitudinal Cohort

| | Variable | Baseline(N=793) | FU1(N=665) | FU2(N=603) | FU3(N=353) |
|---|---|---|---|---|---|
| | Age | 19.07(1.82) | 19.58(1.78) | 20.09(1.87) | 20.50(1.62) |
| Gender | Male | 257(32.4%) | 185(27.8%) | 160(26.5%) | 102(28.9%) |
| | Female | 536(67.6%) | 480(72.2%) | 443(73.5%) | 251(71.1%) |
| Internet use | IAT | 50.82(14.41) | 50.73(13.42) | 50.14(14.11) | 50.97(13.81) |
| | Internet use time | 9.03(2.90) | 8.85(2.85) | 8.81(2.88) | 9.18(2.89) |
| | Internet entertainment time | 6.11(2.76) | 6.08(2.69) | 6.10(4.05) | 6.51(2.79) |
| | QGU-B | 27.59(12.71) | 28.18(12.96) | 27.53(13.07) | 29.69(13.35) |
| | DSM-5 | 3.08(2.41) | 2.65(2.33) | 2.63(2.38) | 2.76(2.47) |
| | IGDS9-SF | 19.18(6.34) | 18.85(6.11) | 18.50(6.04) | 19.03(6.07) |
| Mental health | PHQ-9 | 7.16(4.52) | 7.31(4.30) | 7.36(4.70) | 7.74(4.63) |
| | GAD-7 | 5.46(4.03) | 5.73(3.93) | 5.98(3.82) | 6.46(3.71) |
| | PSS-10 | 30.00(3.88) | 28.83(4.27) | 28.59(4.12) | 28.01(4.70) |
| ASLEC | Interpersonal relationships | 8.07(5.10) | 7.93(5.52) | 7.69(5.37) | 7.90(5.50) |
| | Study pressure | 8.86(4.55) | 7.46(4.68) | 7.29(4.52) | 6.87(4.68) |
| | Being punished | 4.97(5.99) | 4.37(6.16) | 4.02(5.87) | 4.30(5.91) |
| | Bereavement | 2.96(3.51) | 2.76(3.60) | 2.49(3.45) | 2.54(3.51) |
| | Health adaptation | 4.06(2.80) | 4.10(2.94) | 3.87(2.79) | 4.05(2.84) |
| SDQ | Emotional symptoms | 3.25(2.33) | 3.28(2.37) | 3.34(2.33) | 3.62(2.41) |
| | Conduct problems | 2.13(1.12) | 2.17(1.37) | 2.20(1.37) | 2.27(1.34) |
| | Hyperactivity | 4.15(1.39) | 4.22(1.43) | 4.26(1.46) | 4.23(1.49) |
| | Peer problems | 4.51(1.32) | 4.49(1.33) | 4.54(1.33) | 4.44(1.33) |
| | Prosocial | 7.66(1.91) | 7.46(2.02) | 7.31(2.02) | 7.14(2.21) |
| SRQ | Admiration | 24.97(2.38) | 24.93(2.42) | 24.82(2.48) | 24.65(2.60) |
| | Negative social potency | 28.72(4.38) | 28.64(4.52) | 28.62(4.38) | 27.83(4.86) |
| | Passivity | 12.41(3.65) | 12.48(3.58) | 12.04(3.60) | 11.92(3.60) |
| | Prosocial interactions | 29.48(3.04) | 29.29(3.09) | 29.22(3.08) | 29.05(3.10) |
| | Sexual relationships | 9.20(4.54) | 9.41(4.48) | 9.34(4.50) | 9.48(4.50) |
| | Sociability | 13.55(4.01) | 13.17(4.25) | 13.04(4.11) | 12.71(4.13) |

IAT: Internet Addiction Test; QGU-B: Questionnaire of gaming urges–brief; DSM-5: Diagnostic and Statistical Manual of Mental Disorders, Fifth Edition; IGDS9-SF: Internet Gaming Disorder Scale–Short Form; PHQ-9:

Patient Health Questionnaire-9; GAD-7: Generalized Anxiety Disorder 7-item Scale; PSS-10: Perceived Stress Scale–10-item; ASLEC: Adolescent Self-Rating Life Events Checklist; SDQ: Strengths and Difficulties Questionnaire; SRQ: Social Reward Questionnaire.

#### Table S6 Factor Analysis Results and Model Fit Indices for Gaming Motivations

| Parameter | k=2 | k=3 | k=4 | k=5 |
|---|---|---|---|---|
| RMSR | 0.02 | 0.01 | <0.01 | 0 |
| the df corrected root mean square of the residuals | 0.03 | 0.02 | 0.02 | 0 |
| empirical chi square | 105.35 | 24.05 | 4.56 | 0 |
| prob | p < 1.5E-16 | p < 0.0011 | p < 0.1 | |
| liklihood chi square | 58.68 | 15.18 | 2.56 | 0 |
| prob | p < 9E-08 | p < 0.034 | p < 0.28 | |
| TLI | 0.919 | 0.973 | 0.994 | 1.023 |
| RMSEA | 0.028 | 0.016 | 0.008 | 0 |
| BIC | -50.57 | -43.65 | -14.26 | NA |
| fit based upon off diagonal values | 0.96 | 0.99 | 1 | 1 |

RMSR: root mean square of the residuals; TLI: tucker lewis index; RMSEA: root mean square error of approximation; BIC: bayesian information criterion.

#### Table S7 Neuroimaging Data Acquisition

| Variable | | Baseline(N=793) | EEG(N=136) | fMRI(N=91) |
|---|---|---|---|---|
| Age | | 19.07(1.82) | 21.96(2.01) | 21.98(1.93) |
| Sex | Male | 257(32.4%) | 61(44.8%) | 38(41.7%) |
| | Female | 536(67.6%) | 75(55.2%) | 53(58.3%) |
| Internet use | IAT | 50.82(14.41) | 50.94(14.85) | 51.20(15.66) |
| | Internet use time | 9.03(2.90) | 8.85(2.98) | 9.23(2.93) |
| | Internet entertainment time | 6.11(2.76) | 6.27(2.64) | 6.52(2.85) |
| | QGU-B | 27.59(12.71) | 28.63(13.38) | 30.38(13.88) |
| | DSM-5 | 3.08(2.41) | 3.27(2.5) | 2.63(2.38) |

| | | | | |
|---|---|---|---|---|
| | IGDS9-SF | 19.18(6.34) | 19.99(6.39) | 21.12(7.50) |
| Mental health | PHQ-9 | 7.16(4.52) | 7.22(4.57) | 6.90(4.46) |
| | GAD-7 | 5.46(4.03) | 5.61(4.13) | 5.03(3.28) |
| | PSS-10 | 30.00(3.88) | 30.26(4.10) | 29.08(3.90) |
| ASLEC | Interpersonal relationships | 8.07(5.10) | 8.20(5.13) | 8.19(5.63) |
| | Study pressure | 8.86(4.55) | 8.32(4.61) | 8.08(4.56) |
| | Being punished | 4.97(5.99) | 5.23(6.25) | 5.24(7.12) |
| | Bereavement | 2.96(3.51) | 3.28(3.59) | 3.01(4.08) |
| | Health adaptation | 4.06(2.80) | 3.87(2.56) | 4.07(3.26) |
| SDQ | Emotional symptoms | 3.25(2.33) | 3.23(2.30) | 3.43(2.23) |
| | Conduct problems | 2.13(1.12) | 2.33(1.41) | 2.42(1.27) |
| | Hyperactivity | 4.15(1.39) | 4.30(1.38) | 4.44(1.31) |
| | Peer problems | 4.51(1.32) | 4.66(1.40) | 4.32(1.36) |
| | Prosocial | 7.66(1.91) | 7.73(1.72) | 7.42(1.82) |
| SRQ | Admiration | 24.97(2.38) | 24.80(2.60) | 24.59(2.39) |
| | Negative social potency | 28.72(4.38) | 28.73(4.45) | 27.70(4.35) |
| | Passivity | 12.41(3.65) | 12.35(3.69) | 11.55(3.06) |
| | Prosocial interactions | 29.48(3.04) | 29.59(2.91) | 29.29(3.17) |
| | Sexual relationships | 9.20(4.54) | 9.98(4.27) | 10.31(3.83) |
| | Sociability | 13.55(4.01) | 14.09(3.46) | 13.51(3.55) |

IAT: Internet Addiction Test; QGU-B: Questionnaire of gaming urges–brief; DSM-5: Diagnostic and Statistical Manual of Mental Disorders, Fifth Edition; IGDS9-SF: Internet Gaming Disorder Scale–Short Form; PHQ-9: Patient Health Questionnaire-9; GAD-7: Generalized Anxiety Disorder 7-item Scale; PSS-10: Perceived Stress Scale–10-item; ASLEC: Adolescent Self-Rating Life Events Checklist; SDQ: Strengths and Difficulties Questionnaire; SRQ: Social Reward Questionnaire.

### Table S8 Significant Activations for Game and Social Cues

| game_pos_neu | cluster k | T | MNI | | |
|---|---|---|---|---|---|
| | | | x | y | z |
| Pos>Neu | | | | | |
| Lingual_R | 390 | 9.86 | 9 | -82 | -11 |
| Fusiform_L | 226 | 9.56 | -30 | -70 | -17 |
| Precuneus_R | 30 | 7.83 | 21 | -55 | 19 |
| Parietal_Inf_R | 278 | 7.5 | 51 | -46 | 52 |
| Temporal_Mid_R | 165 | 7.02 | 48 | -64 | 10 |
| Frontal_Mid_2_R | 67 | 6.57 | 48 | 41 | 10 |
| Pos<Neu | | | | | |
| Calcarine_L | 610 | 12.19 | -9 | -79 | 4 |
| Fusiform_L | 78 | 9.58 | -30 | -34 | -17 |

| | | | | | |
|---|---|---|---|---|---|
| Frontal_Inf_Orb_2_L | 111 | 9.03 | -30 | 35 | -8 |
| Frontal_Sup_2_L | 165 | 8.47 | -21 | 26 | 40 |
| Frontal_Sup_2_L | 100 | 7.9 | -21 | 59 | 13 |
| Supp_Motor_Area_L | 80 | 6.96 | -12 | 11 | 67 |
| Postcentral_R | 43 | 6.68 | 24 | -28 | 58 |
| Frontal_Inf_Orb_2_R | 23 | 6.43 | 30 | 35 | -8 |
| Frontal_Sup_2_R | 23 | 6.01 | 24 | 35 | 34 |
| Occipital_Mid_L | 31 | 5.96 | -39 | -70 | 28 |
| Heschl_R | 48 | 5.88 | 36 | -28 | 13 |
| game_neg_neu | | | | | |
| Neg>Neu | | | | | |
| Calcarine_R | 400 | 11.97 | 18 | -58 | 16 |
| Lingual_L | 201 | 10.27 | -6 | -79 | -11 |
| Temporal_Mid_R | 921 | 9.97 | 60 | -52 | 16 |
| Temporal_Mid_L | 248 | 9.87 | -60 | -55 | 19 |
| Temporal_Mid_L | 96 | 9.87 | -54 | -22 | -8 |
| Calcarine_L | 29 | 7.52 | -18 | -61 | 16 |
| Temporal_Pole_Sup_L | 32 | 6.74 | -51 | 8 | -23 |
| Frontal_Mid_2_R | 36 | 6.54 | 48 | 17 | 46 |
| Frontal_Inf_Tri_R | 50 | 6.47 | 45 | 11 | 22 |
| Frontal_Inf_Tri_R | 74 | 6.33 | 57 | 29 | 10 |
| Fusiform_R | 22 | 5.93 | 33 | -64 | -14 |
| Neg<Neu | | | | | |
| Precuneus_L | 28 | 11.36 | -6 | -55 | 10 |
| Fusiform_L | 57 | 10.83 | -27 | -34 | -17 |
| Frontal_Sup_Medial_L | 242 | 9.2 | -9 | 59 | 1 |
| Frontal_Sup_2_L | 263 | 9.06 | -21 | 32 | 46 |
| Frontal_Inf_Orb_2_L | 27 | 8.14 | -27 | 35 | -11 |
| Cuneus_L | 121 | 8.03 | -6 | -88 | 16 |
| Heschl_R | 107 | 7.25 | 51 | -7 | 4 |
| social_pos_neu | | | | | |
| Pos>Neu | | | | | |
| Lingual_R | 528 | 12.14 | 9 | -82 | -2 |
| Precuneus_L | 193 | 9.62 | -3 | -55 | 22 |
| Fusiform_L | 320 | 9.55 | -39 | -76 | -17 |
| Precentral_L | 331 | 8.59 | -36 | -22 | 58 |
| Supp_Motor_Area_L | 62 | 7.75 | -6 | -4 | 58 |
| Temporal_Sup_L | 33 | 7 | -60 | -1 | -8 |

| | | | | | |
|---|---|---|---|---|---|
| Rolandic_Oper_L | 78 | 6.63 | -45 | -25 | 19 |
| Frontal_Sup_Medial_L | 27 | 5.82 | -6 | 65 | 7 |
| Pos<Neu | | | | | |
| Frontal_Inf_Tri_R | 589 | 10.07 | 42 | 32 | 16 |
| Frontal_Inf_Orb_2_L | 494 | 9.58 | -45 | 38 | -5 |
| Fusiform_L | 54 | 7.25 | -27 | -43 | -14 |
| Fusiform_R | 57 | 7.19 | 30 | -40 | -14 |
| Occipital_Mid_R | 28 | 7 | 39 | -73 | 28 |
| Parietal_Inf_L | 120 | 6.99 | -57 | -49 | 37 |
| Precuneus_R | 25 | 6.94 | 18 | -55 | 16 |
| Supp_Motor_Area_L | 219 | 6.93 | -9 | 17 | 67 |
| Temporal_Inf_L | 102 | 6.74 | -57 | -55 | -11 |
| Temporal_Inf_R | 61 | 6.01 | 51 | -43 | -17 |
| Frontal_Mid_2_L | 57 | 5.78 | -39 | 26 | 43 |
| social_neg_neu | | | | | |
| Neg>Neu | | | | | |
| Temporal_Mid_L | 953 | 12.63 | -45 | -64 | 7 |
| Lingual_L | 401 | 12.53 | -6 | -73 | 1 |
| Occipital_Mid_R | 620 | 9.62 | 51 | -76 | 1 |
| Occipital_Sup_L | 82 | 8.76 | -18 | -88 | 25 |
| Fusiform_L | 23 | 8.07 | -42 | -49 | -23 |
| Postcentral_L | 412 | 7.65 | -48 | -16 | 49 |
| Supp_Motor_Area_L | 75 | 6.83 | -6 | 2 | 49 |
| Precuneus_R | 49 | 6.79 | 12 | -64 | 55 |
| Frontal_Sup_Medial_L | 52 | 6.39 | 6 | 56 | 25 |
| Frontal_Sup_2_R | 37 | 6.37 | 24 | -4 | 52 |
| Neg<Neu | | | | | |
| Fusiform_L | 167 | 15.83 | -30 | -49 | -14 |
| Precuneus_L | 120 | 14.93 | -9 | -55 | 10 |
| Fusiform_R | 390 | 14.78 | 27 | -40 | -17 |
| Precuneus_R | 73 | 13.51 | 12 | -55 | 13 |
| Cingulate_Mid_L | 196 | 13.46 | 0 | -34 | 37 |
| Frontal_Inf_Tri_R | 233 | 10.87 | 45 | 38 | 13 |
| Occipital_Mid_L | 479 | 10.17 | -36 | -85 | 19 |
| Frontal_Inf_Orb_2_L | 20 | 10.06 | -30 | 35 | -11 |

| | | | | | |
|---|---|---|---|---|---|
| Angular_R | 332 | 9.5 | 42 | -67 | 31 |
| Temporal_Inf_L | 67 | 9.31 | -57 | -49 | -14 |
| Temporal_Mid_R | 57 | 8.99 | 60 | -40 | -11 |
| Frontal_Sup_2_R | 143 | 7.97 | 24 | 23 | 52 |
| Frontal_Sup_2_L | 243 | 7.67 | -24 | 20 | 52 |
| Frontal_Mid_2_L | 69 | 7.39 | -45 | 35 | 19 |
| Frontal_Med_Orb_R | 36 | 7.18 | 3 | 50 | -5 |

A whole-brain voxel-wise family-wise error (FWE) correction was applied, with the statistical significance set at $p < 0.05$. Pos = positive cues; Neg = negative cues; Neu = neutral cues.

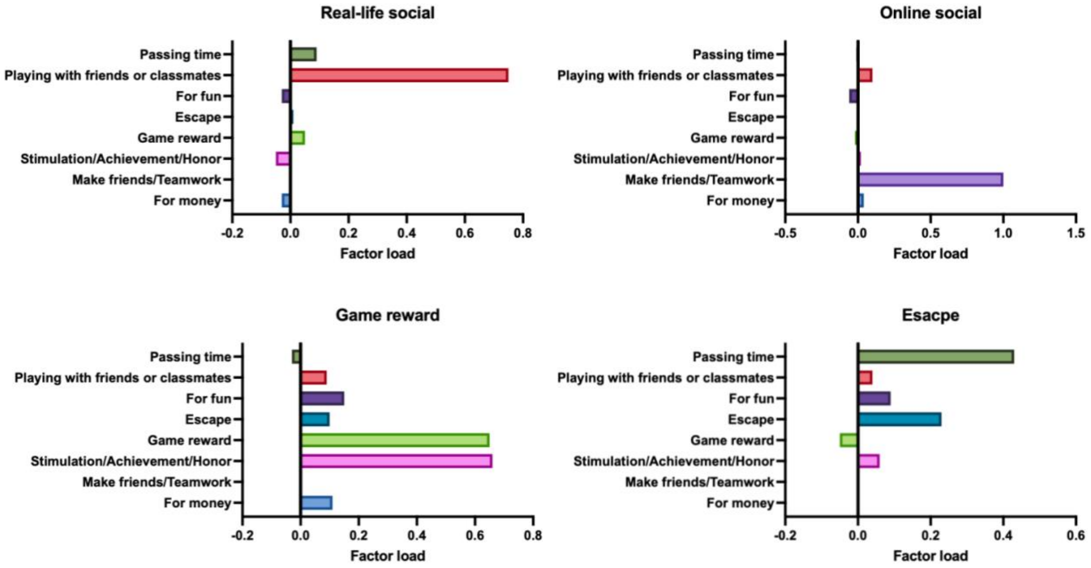


**Figure S1 Factor Loadings of Gaming Motivation Items in the Longitudinal Sample** Four parts show the factor loadings for real-life social, online social, game reward, and escape motivations, respectively, derived from the overall sample (N=793).
